# Untargeted metabolomics reveals a reproducible signature of fatal insulin intoxication

**DOI:** 10.64898/2026.02.27.26347264

**Authors:** Albert Elmsjö, Carl Söderberg, Fredrik Tamsen, Henrik Green, Fredrik C. Kugelberg, Liam J. Ward

**Author notes:** Corresponding author: Liam J. Ward, *PhD, Associate Professor.

## Abstract

Fatal insulin intoxication remains difficult to diagnose because insulin rapidly degrades after death, limiting the reliability of direct biochemical measurements. We hypothesised that insulin excess induces systemic metabolic alterations that persist beyond insulin degradation and can be captured using postmortem metabolomics. High-resolution mass spectrometry (HRMS)-based metabolomics was applied to a national cohort (2017-2024), including 51 fatal insulin intoxications and non-insulin deaths for comparison. Orthogonal partial least squares-discriminant analysis (OPLS-DA) models were trained on cases collected between 2017-2022. Performance was evaluated using two temporally distinct test sets (2023-2024), a matched validation cohort (n=59) and a heterogeneous cohort (n=154), with 14 insulin cases included. An insulin-associated metabolomic signature comprising 91 features demonstrated reproducible discrimination. In the matched cohort, insulin intoxication classification achieved 100% sensitivity and 73% specificity. In the heterogeneous cohort, 100% sensitivity was maintained with 72% specificity. No Insulin false negatives were observed in either test set. Significant alterations were observed across acylcarnitines, fatty acids/lipids, and purine/nucleoside metabolites. Fatal insulin intoxication is associated with a reproducible metabolomic signature. While not suitable as a confirmatory diagnostic approach, the high sensitivity and absence of false negatives suggests potential utility in supporting exclusion of insulin intoxication in death investigations.

## INTRODUCTION

Insulin intoxication remains one of the most challenging causes of death to diagnose, largely due to the biochemical instability of insulin after death. These challenges have been recognised for decades and arise from the rapid degradation of insulin and the difficulty of distinguishing exogenous administration from endogenous insulin postmortem. Such intoxications may arise from accidental overdose, suicide, or homicide, but postmortem confirmation is often difficult [1]. Insulin degrades rapidly after death, analytical sensitivity is limited, and interpretation is further complicated by postmortem redistribution, haemolysis, and medical intervention prior to death [2]. Importantly, absence of detectable insulin does not necessarily exclude insulin-induced hypoglycaemia. As a result, confirmation of insulin intoxication may rely on circumstantial evidence rather than definitive toxicological findings, and some cases may remain undetected, raising concern that the true incidence rate of insulin intoxication is underestimated [1,3].

The diagnostic difficulty associated with insulin intoxication is closely linked to its mechanism of action. Excess insulin leads to hypoglycaemia, resulting in a cascade of downstream metabolic disturbances affecting cellular energy metabolism and substrate utilisation [4]. Whilst insulin itself may no longer be detectable postmortem, the downstream metabolic disturbances resulting from hypoglycaemia may persist and remain detectable in postmortem samples. As such, approaches that rely primarily on the direct detection of exogenous compounds are poorly suited to identify such indirect effects, particularly when the causative compound is unstable or rapidly degraded.

Postmortem metabolomics offers an alternative strategy by enabling comprehensive profiling of endogenous metabolites that reflect the metabolic state immediately preceding death. Rather than targeting a single compound, metabolomic approaches assess multivariate patterns that may be characteristic of specific causes of death. In recent years, postmortem metabolomics has demonstrated potential in large-scale cause of death screening [5], and in the discrimination of specific fatal conditions, including pneumonia, opioid intoxications, and hypothermia [6–9]. These findings suggest that metabolomic signatures may remain detectable in postmortem material, even when conventional toxicological targets are absent or inconclusive.

In a previous study [10], we applied postmortem metabolomics to a cohort of confirmed insulin intoxication and identified a distinct metabolomic signature that discriminated these deaths from hyperglycaemic diabetic coma cases and presumed normoglycaemic control cases. That study demonstrated clear group separation and identified a metabolomic signature including reductions in multiple hydroxylated-acylcarnitines, consistent with an altered energy metabolism in the context of excess insulin activity. When applied as a screening tool in a large set of autopsy cases, this signature highlighted a small number of deaths with plausible hypoglycaemia-related mechanisms. These findings provided an important proof-of-concept [10], but the study was limited by the number of insulin intoxication cases available and did not include independent validation using temporally distinct cases.

Before postmortem metabolomic approaches can be adopted in routine forensic practice, their analytical stability and reproducibility must be established. Currently, it remains unclear whether metabolomic signatures derived from retrospective insulin intoxication cases retain discriminatory capacity when applied to independent cases collected at later time points, and whether such signatures persist across heterogeneous populations representative of routine practice. Addressing these issues is essential to discriminate true biological signal from model overfitting, and to evaluate the robustness and reproducibility of an insulin-associated metabolomic signature, and explore the potential role as complementary biochemical evidence in forensic death investigations where direct insulin measurements are unreliable.

In the present study, we expand upon our previous work by substantially increasing the number of confirmed insulin intoxications, and by employing a temporally separated training and validation framework across different acquisition years. Using postmortem high-resolution mass spectrometry (HRMS) data generated from routine toxicological screening between 2017-2024, models were developed from 2017-2022 and subsequently evaluated on temporally distinct cases from 2023-2024. Model performance was assessed under two complementary validation conditions: a controlled test set matched for demographics, and a second test set designed to represent a heterogeneous forensic cohort without restrictions. The aim of this study was to evaluate the robustness and reproducibility of an insulin-associated metabolomic signature detectable postmortem, and its potential utility as a biologically grounded data-driven support tool for death investigations.

## METHODS

The human study was approved by the Swedish Ethical Review Authority (Dnr 2019-04530, 2024-05087-02, 2025-02459-02). Due to the retrospective nature of the study, the need for informed consent was waived by the Swedish Ethical Review Authority. All methods were carried out in accordance with relevant guidelines and regulations.

### Study population

Human postmortem cases admitted to the National Board of Forensic Medicine, Sweden, between July 2017 and December 2024 were considered for inclusion in the study (*n* = 40,595; Figure 1). This cohort represents the entire Swedish forensic system and includes cases from all six regional Departments of Forensic Medicine. Toxicological screening is performed on the majority of forensic autopsy cases (>95%), and all biological samples are analysed centrally at the national laboratory at the Department of Forensic Genetics and Forensic Toxicology, Linköping, Sweden. Since July 2017, all high-resolution mass spectrometry (HRMS) data generated during routine toxicological screening of autopsy cases have been collated in a database, and is available for retrospective analysis in postmortem metabolomics investigations.

**Figure 1:**
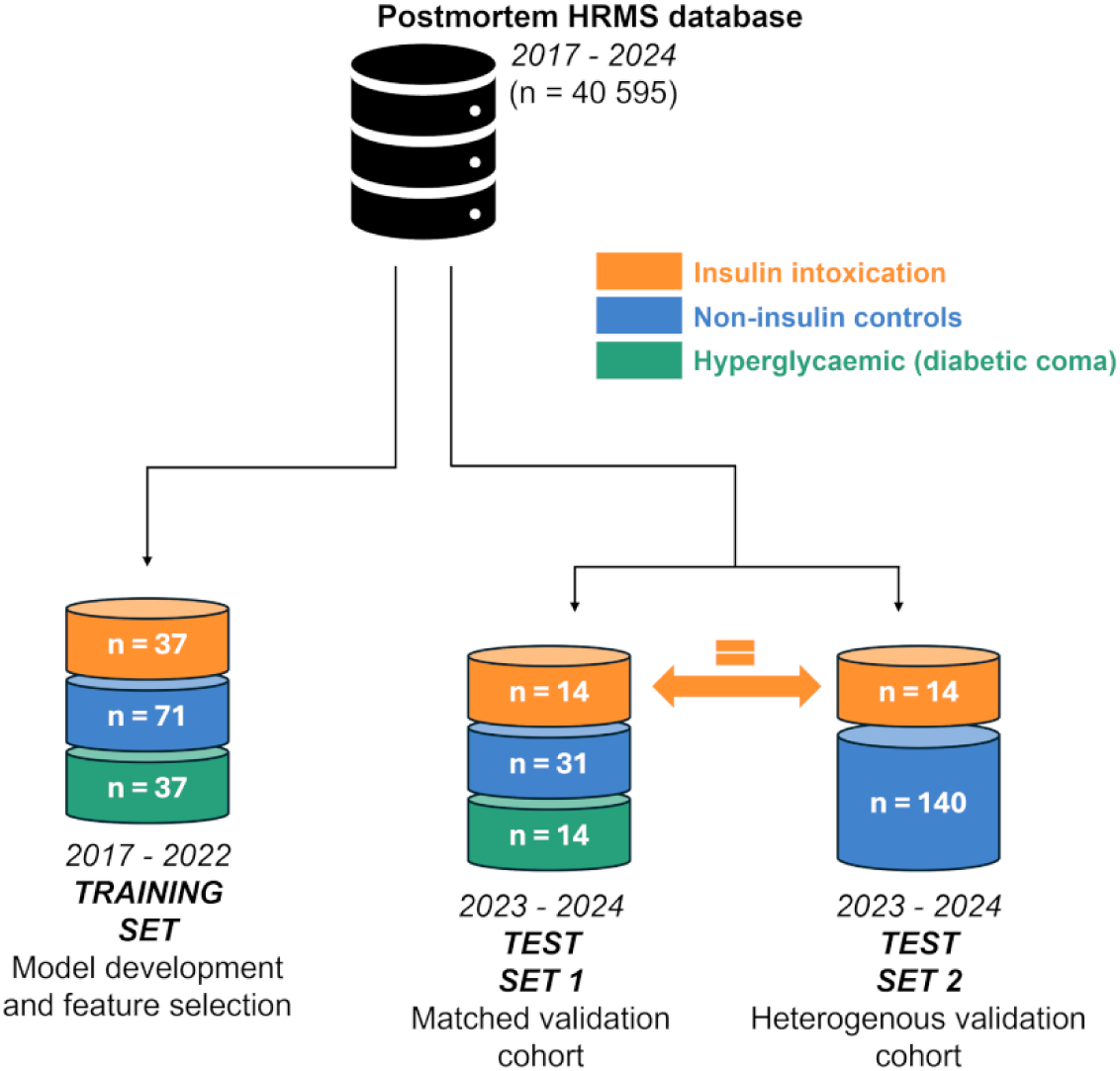
Study design and cohort allocation. Cases from the HRMS database of Swedish forensic autopsy cases (2017–2024; n = 40,595) were eligible for inclusion, all confirmed insulin intoxication deaths (n = 51) were identified and split temporally into a training cohort (2017–2022, n = 37) and an independent test cohort (2023–2024, n = 14). Training controls comprised hanging deaths (n = 71) and hyperglycaemic diabetic coma deaths (n = 37). Two temporally distinct validation sets were constructed: a matched three-group validation cohort (Test Set 1) and a heterogeneous forensic validation cohort (Test Set 2). Numbers denote cases per group.

From this database, all autopsy cases in which the cause of death has been diagnosed as insulin intoxication by a forensic pathologist were identified (n = 51). No cases were excluded, as all cases were >17 years old and had available HRMS data. These cases were stratified by calendar year into a training cohort of deaths between 2017 and 2022 (n = 37), and a test cohort of deaths between 2023 and 2024 (n = 14). All cases are considered confirmed insulin intoxications. Confirmation was based on the forensic pathologists’ cause-of-death determination, supported by analytical findings where available and/or strong circumstantial evidence (e.g. scene findings, medical history).

For the training set, additional comparison groups were selected from the same period (2017–2022). Two groups were selected to represent deaths where the decedent is assumed to be hyperglycaemic and normoglycaemic to contrast the hypoglycaemic state of insulin intoxication. These deaths were diagnosed as diabetic coma (n = 37), hereby referred to as the hyperglycaemic or “hyper” group, and hanging (n = 71) representing the “control” group of presumed normoglycaemic deaths. These groups were selected to enable a structured comparison of metabolomic profiles across deaths associated with low, normal, and high glycaemic states.

For the test period (2023–2024), two test sets were constructed. The first (Test Set 1) mirrored the design of the training cohort and consisted of insulin intoxications (n = 14), hyperglycaemic deaths (n = 14, diabetic coma), and controls (n = 31, hanging). The second (Test Set 2) was designed to reflect forensic casework heterogeneity and included the same insulin intoxications (n = 14) together with a random selection of autopsy cases (n = 140) without restrictions on cause of death diagnosis. Due to the limited number of insulin intoxication in the test period (2023-2024), it was decided that the same positive cases would be used in both test sets and evaluation based on performance within increased heterogeneity of negative cases in Test Set 2. Overall, this design allowed the evaluation of constructed models under controlled, matched conditions (Test Set 1), but also in a heterogenous population (Test Set 2) that more closely reflects routine forensic practice testing performance.

For both training set and Test Set 1, cases were matched as closely as possible on age, sex, body mass index (BMI), and postmortem interval (PMI) to minimise confounding effects on the metabolomic profiles. For Test Set 2, random cases were frequency-matched by sex to insulin intoxication cases to account for the male predominance in the general forensic autopsy population. The difference in sex distribution between the insulin cohort and the general forensic autopsy population is 60% males and 73% males, respectively. Test Set 2 was intended to test model performance under heterogenous operational conditions.

### Samples preparation and HRMS acquisition

Data was obtained routine toxicological screening of postmortem femoral whole blood samples from autopsies conducted at the National Board of Forensic Medicine, Sweden. Routine toxicological screening is performed using HRMS, and resulting data were retrospectively extracted for metabolomics data.

Sample preparation and HRMS acquisition were performed as part of the accredited routine toxicological workflow and the methods described in detail previously [23]. Briefly, samples were subjected to a protein precipitation using organic solvent containing internal standards, followed by centrifugation and analysis of the supernatant by liquid chromatography-quadrupole time-of-flight mass spectrometry (LC-QTOF-MS) operated under standardised conditions. Sample separation was performed using gradient elution on a C18 column (150 mm x 2.1 mm, 1.8 µm; Water Acquity HSS T3 column). Samples admitted between July 2017 and November 2020 were run on an Agilent 6540 QTOF system in MS-mode only, and samples admitted after November 2020 were run on an Agilent 6546 QTOF system with data-dependent acquisition (auto-MSMS). All source settings and TOF parameters (gas temperatures, voltages, and mass ranges) were consistent between QTOF systems.

Raw HRMS data files were converted to open-source formats using msConvert [24] before processing using XCMS for feature detection, retention time alignment, and peak grouping. Chromatographic features were defined by mass-to-charge ratio (m/z) and retention time and exported as a feature matrix for further analysis. Data were normalised using probabilistic quotient normalisation, followed by log transformation prior to multivariate data analysis using SIMCA.

### Model construction

Multivariate modelling was performed using orthogonal partial least squares discriminant analysis (OPLS-DA) implemented in SIMCA. Model construction was limited to just the training cohort (2017-2022) to avoid information leakage into the test datasets.

To identify metabolite features specifically associated with insulin intoxication, we conducted OPLS-DA modelling within a step wise framework. First, we build two OPLS-DA models comparing insulin intoxications with the control groups, and insulin intoxications with hyper group. Model performance was evaluated using the explained variance (R^2^) and the predictive ability (Q^2^) derived from a seven-fold cross validation. Next features contributing to discrimination, with a variable influence on projection (VIP) > 1, in either OPLS-DA models were combined into a shared-and-unique-structures (SUS) plot. Features consistent with insulin intoxication, exceeding p(corr) thresholds of −0.2 and 0.2 along both plot axes, were identified and extracted for further modelling. The final OPLS-DA model included only features identified to discriminate insulin intoxications from the SUS-plot and these were plotted in a three-group OPLS-DA model and model discrimination evaluated using R2 and Q2. For all constructed OPLS-DA models, permutation testing was performed cycling 500 random class permutations to test for any model overfitting.

### Test set evaluation

For both Test Set 1 and Test Set 2, class prediction was performed using the final OPLS-DA model in SIMCA, built from the 91 insulin-associated features. Prior to classification, model applicability was assessed using distance to model in X-space (DModX). Samples exceeding the predefined critical limit (α = 0.01; 99% confidence), were excluded to restrict predictions to the model space.

For Test Set 1, the final three-group OPLS-DA model was used to classify cases as control, hyper, or insulin. Predictions were then compared with forensic cause of death diagnoses using a confusion matrix, and performance was evaluated via sensitivity, specificity, accuracy, and receiver-operating-characteristic (ROC) analysis.

For Test Set 2, the model was reformulated as a binary OPLS-DA model, where control and hyper cases were combined into a single non-insulin group modelled against insulin intoxications. Cases within the applicability domain were classified as insulin or non-insulin and evaluated against true diagnoses using a confusion matrix. Predictive performance was assessed using sensitivity, specificity, accuracy, and ROC analysis.

### Metabolite identification

Metabolite feature identification was performed via matching feature m/z values and retention times (rt) to reference metabolites included in the internal metabolite library at the National Board of Forensic Medicine, Sweden. If a feature was not present in the internal metabolite library, then the feature m/z was matched to online public databases, such as the human metabolome database (HMDB), and METLIN databases. In accordance with the Metabolomics Standards Initiative (MSI) for feature annotation, such identification represent levels 2 putative metabolite identification [17].

### Univariate statistical analysis and effect size estimation

To further characterise the metabolites of the insulin-signature from the SUS analysis, all annotated features were evaluated across the full cohort irrespective of the multivariate modelling framework. Analyses were restricted to insulin intoxication cases and non-insulin cases (control and random cohorts), with hyper cases excluded to avoid any potential confounding effect by opposing glycaemic states. PQN-normalised feature intensities were log2-transformed prior statistical testing. For each metabolite, between group differences were assessed using Wilcoxon rank-sum tests. Resulting p-values were adjusted for multiple testing using Benjamini-Hochberg false discovery rate (FDR). Log2 fold changes were calculated as the difference in median log2 intensities between insulin intoxication and non-insulin groups.

To quantify the magnitude of the group separation independent of statistical significance, effect sizes were estimated using Cliff’s delta, a non-parametric rank-based measure of stochastic superiority. Cliff’s delta ranges from −1 to +1, where negative values indicate an increased abundance in insulin cases and positive values indicate decreased abundances.

Volcano plots, effect-size forest plots, and box-and-jitter visualisations were generated in R (v4.4.2) using *ggplot2*, with additional packages *ggrepel, ggpubr,* and *effsize* for annotation and effect size estimation.

## RESULTS

### Cohort characteristics

Study population, cohort selection, and temporal division are presenting in Figure 1. A demographic overview of the forensic autopsy cases selected for inclusion in the training set, Test Set 1, and Test Set 2, are presented in Table 1. Insulin intoxication, hyperglycaemic deaths (referred to as “hyper”), and control deaths included for the training set and Test Set 1 were matched for sex, age, BMI and PMI and no significant differences were observed within these groups. Test Set 2 was designed to reflect routine forensic casework heterogeneity and comprised of insulin intoxication cases and randomly selected autopsy cases. No statistically significant differences in age, BMI or PMI were observed between groups in Test Set 2. Insulin intoxication cases included in Test Set 1 and Test Set 2 were identical.

**Table 1:** Case demographics.

|  | **Training set (2017-22)<sup>†</sup>** | **Test Set 1 (2023-24)<sup>†</sup>** | **Test Set 2 (2023-24)<sup>‡</sup>** |
| --- | --- | --- | --- |
| **Insulin intox.** | n = 37 | n = 14<sup>§</sup> | n = 14<sup>§</sup> |
| **Hyperglycaemic** | n = 37 | n = 14 | - |
| **Control (hanging)** | n = 71 | n = 31 | - |
| **Random cases** | - | - | n = 140 |
| **Sex, (F / M) %** | 40 / 60 | 44 / 56 | 43 / 57 |
| **Age, years** | 59 (42 – 69) | 58 (43 – 70) | 60 (48 – 71) |
| **BMI, kg/m<sup>2</sup>** | 25.0 ( 21.9 – 28.8) | 26.0 (23.5 – 29.2) | 25.6 (22.1 – 28.8) |
| **PMI, days** | 6 (4 – 8) | 5 (4 – 7) | 5 (3 – 7) |
| Data shown as median (Q1-Q3) for continuous variables and percentage for categorical variables |  |  |  |
| † Insulin intoxication, hyperglycaemic, and control cases in training set and Test Set 1 were matched for age, sex, body mass index (BMI), and postmortem interval (PMI), and no statistically significant differences were observed between groups. |  |  |  |
| ‡ Random cases selected in Test Set 2 were frequency-matched on sex to account for the male predominance in the general forensic autopsy population, and no statistically significant differences were observed between groups ($p > 0.05$). | | | |
| § Insulin intoxications cases included in Test Set 1 and Test Set 2 were identical. |  |  |  |

### Discrimination and prediction of insulin intoxication cases

The training set data, 2017–2022, were used to construct two primary orthogonal partial least squares-discriminant analysis (OPLS-DA) models for the discrimination of insulin deaths versus hyper and control deaths. The insulin versus control model was described using 1904 features, and achieved a R2 = 0.91, Q2 = 0.70, and CV-ANOVA *p* = 8.8 × 10^−22^ (Fig. 2A). The insulin versus hyper model was described with 1443 features, and achieved a R2 = 0.90, Q2 = 0.85, and CV-ANOVA *p* = 3.9 × 10^−27^ (Fig. 2B). Permutation tests, cycling 500 permutations, revealed no overfitting of either models (Supplement Figure S1). Further feature reduction was conducted to extract insulin death related features by constructing a shared-and-unique structures (SUS) plot using the two OPLS-DA models, herein, 94 features exceeded set p(corr) thresholds (Fig. 2C). A three group OPLS-DA model was reconstructing using only the remaining 94 features to ensure group separation, this model achieved a R2 = 0.53, Q2 = 0.42, and CV-ANOVA *p* = 1.7 × 10^−22^ (Fig. 2D). Additional permutation tests revealed no overfitting (Supplement Figure S1).

**Figure 2:**
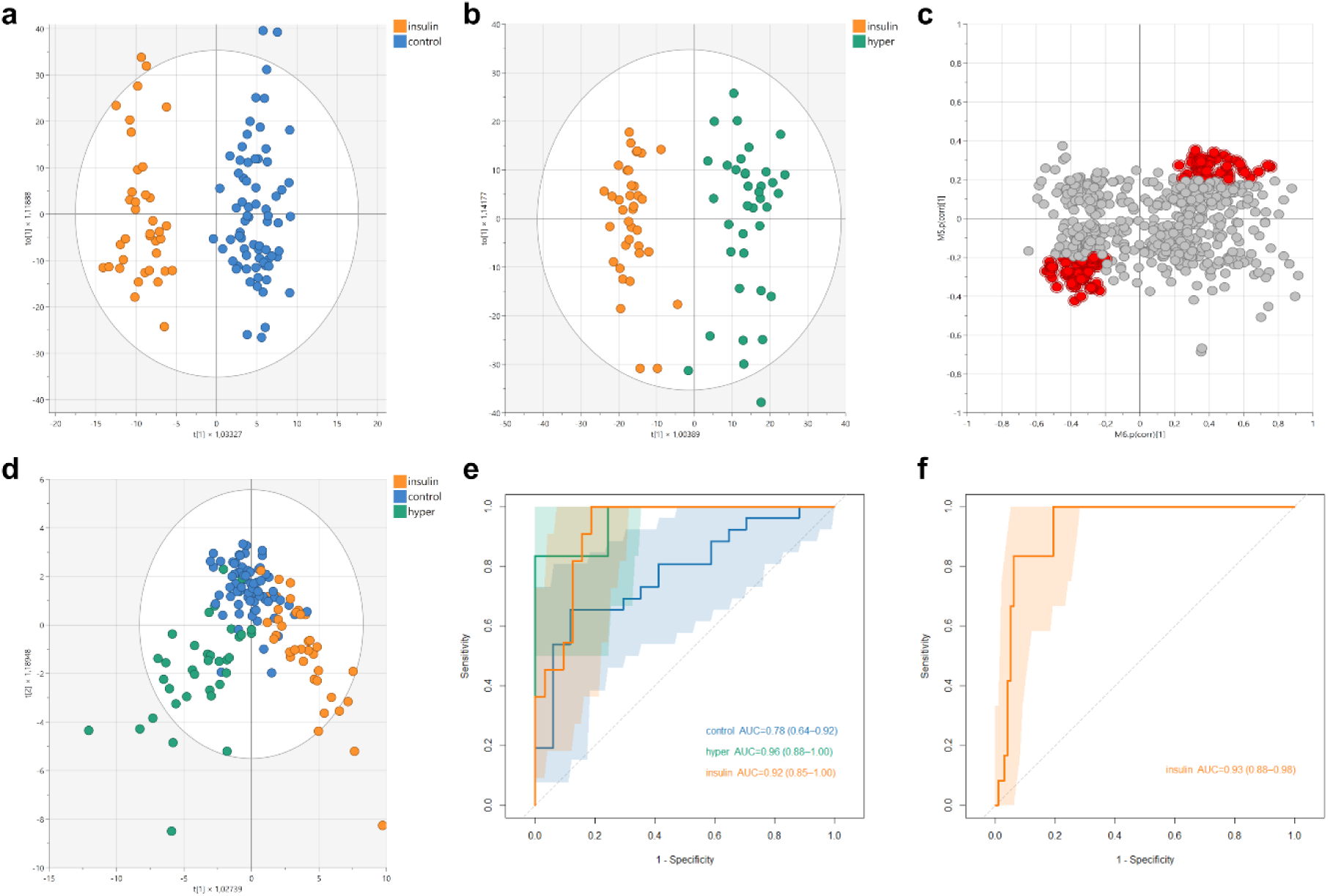
Multivariate modelling, feature extraction, and independent validation of insulin-associated metabolomic signatures. **(a)** Orthogonal partial least squares–discriminant analysis (OPLS-DA) score plot comparing insulin intoxication (n = 37) and control (n = 71) samples in the training cohort, demonstrating separation between groups. **(b)** OPLS-DA score plot comparing insulin intoxication (n = 37) and hyperglycaemic (n = 37) cases, showing distinct clustering. **(c)** Shared-and-unique structures (SUS) plot derived from the two OPLS-DA models, highlighting features (red; n = 91) uniquely associated with insulin intoxication used for feature selection. **(d)** Three-class OPLS-DA model (insulin, n = 37; control, n = 71; hyperglycaemic, n = 37) constructed using the 91 insulin-associated metabolites identified from the SUS analysis, showing preserved class separation with the reduced feature panel. **(e)** Receiver operating characteristic (ROC) curves for validation of class predictions in Test Set 1 (matched cohort; insulin, n = 14; hyper, n = 14; control n = 31). Prior to classification, cases exceeding the predefined distance to model in X-space (DModX) threshold (α = 0.01) were excluded. Performance is summarised by area under the curve (AUC) and 95% confidence intervals (CI). **(f)** ROC curve for validation of insulin intoxication prediction in Test Set 2 (heterogenous cohort; insulin, n = 14; random, n = 140). Prior to classification, cases exceeding the predefined DModX threshold (α = 0.01) were excluded. Performance is summarised by area under the curve (AUC) and 95% confidence intervals (CI).

Receiver operating characteristic (ROC) analysis was performed for both independent Test Set 1, the controlled multiclass cohort, and Test Set 2, the binary cohort representing forensic casework heterogeneity. For both test sets, the applicable domain was established using a distance to model in X-space (DModX) filter with a predefined critical limit (α = 0.01). Cases exceeding this threshold were excluded prior to classification as a quality control measure to restrict predictions to the validated model space and avoid extrapolation beyond the underlying training distribution. In Test Set 1, 16 cases (27.1%), including three insulin intoxications, were excluded; and in Test Set 2, 44 cases (28.6%), including two insulin intoxications, were excluded (Table 2). ROC analysis demonstrated preserved discriminatory performance for insulin intoxication in both validation scenarios, with area under the curve (AUC) values of 0.92 for Test Set 1 (Figure 2E) and 0.93 for Test Set 2 (Figure 2F). In the multiclass setting of Test Set 1, control had an AUC = 0.78 and hyper cases an AUC = 096 (Figure 2E).

**Table 2:** Validation performance in temporally distinct test sets.

|  | Test Set 1 | Test Set 2 |
| --- | --- | --- |
| **Total cases, *n*** | 59 | 154 |
| **Rejected by DModX, *n* (insulin cases)** | 16 (3) | 44 (2) |
| **Coverage, (insulin cases)** | 72.9 % | 71.4 % |
| **Sensitivity – insulin** | 100 % | 100 % |
| **Specificity – insulin** | 73.3 % | 72.5 % |
| **PPV – insulin** | 57.9 % | 30.8 % |
| **NPV – insulin** | 100 % | 100 % |
| **Accuracy** | 76.7 % | 75.5 % |
| Test Set 1: matched validation cohort reflecting the training set design; Test Set 2: heterogenous validation cohort including insulin cases and randomly selected forensic autopsy cases. Sensitivity, specificity, positive predictive value (PPV), negative predictive value (NPV), and accuracy calculated among cases within the distance to model in X-space (DModX)-defined applicability domain. |  |  |

Table 2 presents validation performance within the DModX-defined applicable domain. In Test Set 1, the model achieved 100% sensitivity for insulin intoxication, with 73.3% specificity and an overall accuracy of 76.7%. Predictive performance for hyper and control cases were lower (Supplement Table S1). In Test Set 2, performances were comparable, with insulin intoxication classification retaining 100% sensitivity, with 72.5% specificity and an overall accuracy of 75.5%. Notably, no false negatives were observed within the applicability domain in either test set, resulting in a negative predictive value (NPV) of 100%, whereas the positive prediction value (PPV) was lower in the heterogenous cohort, reflecting the underlying class distribution. Importantly, insulin intoxication cases were still identified with 100% sensitivity in both validation cohorts even when DModX-filtering was not applied, although specificity an overall accuracy were reduced (Supplement Table S2).

### Metabolomic signature

Metabolite annotation was performed for the 91 chromatographic features identified as insulin-associated in the SUS analysis (Supplement Table S3). These metabolites were subsequently evaluated across the full cohort to characterise the metabolomic signature of insulin intoxication. Comparison of all insulin intoxication cases (n = 51) with non-insulin cases (n = 211) demonstrated widespread and statistically significant differences across the insulin-associated metabolite panel (Figure 3A). Several metabolites showed large absolute log_2_ fold changes with strong false discovery rate (FDR)-adjusted significance, indicating robust alterations associated with insulin intoxication. The highlighted identified metabolite features spanned numerous biochemical classes, including acylcarnitines, amino acids/peptides, fatty acids/lipids, and purine/nucleoside metabolites.

**Figure 3:**
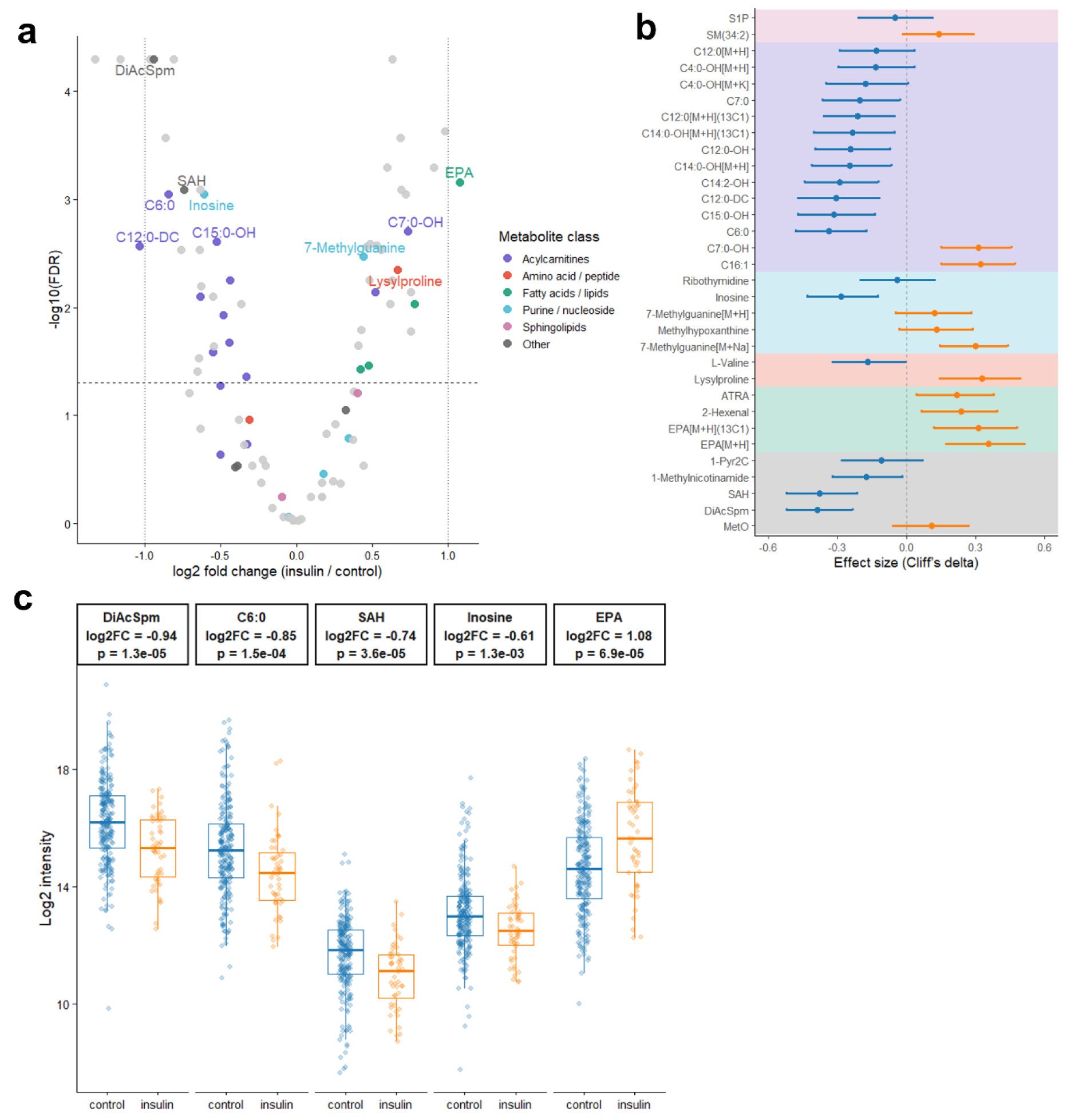
Differences in insulin-associated metabolites. **(a)** Volcano plot showing log₂ fold change versus FDR-adjusted significance for the 91 insulin-signature metabolites identified by SUS analysis, comparing all insulin cases (n = 51) with all non-insulin cases (n = 211; control + random cohort). Metabolites are coloured by biochemical class, and the 10 most discriminatory features are labelled. **(b)** Effect size analysis of the identified insulin-signature metabolites using Cliff ‘s delta with 95% confidence intervals. Positive values indicate higher abundance in insulin intoxication, and negative values indicate lower abundance. Background shading denotes metabolite class. **(c)** Box-and-jitter plots showing log2 intensities for the top five metabolites across groups. Boxes indicate the median and interquartile range, and points represent individual samples. Two-sided Wilcoxon rank-sum test p-values are shown. Analyses were performed on the full cohort, excluding hyperglycaemic cases, and are independent of the SIMCA training/test splits used for predictive modelling. Abbreviations: 1-Pyr3c – 1-Pyrroline-2-carboxylate; ATRA – all-trans-Retinoic acid; C4:0-OH – 3-Hydroxybutryrlcarnitine; C6:0 – Hexanoylcarnitine; C7:0 – Heptanoylcarnitine; C7:0-OH – Hydroxyheptanoylcarnitine; C12:0 – Dodecanoylcarnitine; C12:0-DC – Dodecanedioylcarnitine; C14:0-OH – 3-Hydroxytetradecanoylcarnitine; C14:2-OH – 3-Hydroxy-5,8-tetradecadienoylcarnitine; C15:0-OH – 3-Hydroxypentadecanoyl carnitine; C16:1 – Cervonyl carnitine; DiAcSpm – N1,N12-Diacetylspermine; EPA – Eicosapentaenoic acid; MetO – Methionine sulphoxide; S1P – Sphinganine 1-phosphate; SAH – S-Adenosylhomocysteine; SM(34:2) – Sphingomyelin 34:2.

To quantify the magnitude of and direction of the group separation independent of sample size, non-parametric effect sizes were estimated using Cliff’s delta (Figure 3B). Many metabolites demonstrated moderate effects (δ ∼0.3), indicating separation between insulin and non-insulin cases at the individual level. Effect directions were largely consistent within metabolite classes, with coordinated shifts observed across multiple acylcarnitines and lipid-related metabolites. These findings demonstrate that the differences were not solely driven by statistical power, but reflected biologically meaningful changes in metabolite abundance.

Box-and-jitter visualisation of the top five ranked metabolites confirmed differences in abundance in log_2_ intensities between groups (Figure 3), with all comparisons remaining significant using two-sided Wilcoxon rank-sum testing. These difference were consistent across individuals and independent of the training and test set splits used to multivariate modelling.

## DISCUSSION

In this study, we demonstrate that insulin intoxication is associated with a reproducible metabolomic signature detectable using untargeted HRMS-based metabolomics. Across temporally distinct validation and heterogeneous postmortem populations, the insulin-associated signature retained discriminatory performance, indicating that systemic metabolic consequences of insulin excess persist and can be captured analytically even when direct insulin measurement is challenging. These findings suggest that postmortem metabolomics may provide complementary biochemical evidence in forensic investigations where direct insulin measurements are unreliable.

The main aim of the study was to evaluate whether an insulin-associated metabolomic signature derived from retrospective cases would retain reproducibility when applied to temporally distinct postmortem cases. To address this, models were trained on cases from 2017 – 2022 and evaluated using independent autopsy cases from 2023 – 2024. This temporal separation reduces the risk of data leakage and provides a more rigorous assessment of generalisability than internal cross-validation alone [11]. Performance was assessed under two complementary validation conditions: a matched cohort (Test Set 1) enabling controlled comparison between glycaemic states, and a heterogeneous cohort (Test Set 2) designed to reflect forensic casework with broad biological and pathological variability. This second validation condition provided a more stringent assessment of whether the insulin-associated metabolomic signal remained detectable outside the controlled structure of the training cohort. Within the applicability domain, no false negatives were observed in either test set, resulting in an NPV of 100%. Despite the increased biological variance inherent to these populations, the insulin-associated signature remained stable across acquisition years, supporting the presence of a reproducible biological signal rather than cohort- or model-specific artefacts.

Compared to our earlier proof-of-concept study [10] the expanded cohort increased the number of confirmed insulin intoxications, and introduced a temporal distinct validation. The previous work focussed on the retrospective discrimination within a limited cohort, whereas the current study extends these findings by demonstrating preserved discriminatory performance in later-acquired cases. Importantly, several metabolite classes and individual metabolites identified previously were observed in the present study. Metabolite classes consistent in both studies include acylcarnitines, amino acid / peptides, and purines / nucleotides. Looking at the exact overlap of individual metabolites 30% of the previously identified signature were also identified in the current study, including hydroxylated acylcarnitines (C4-OH, C12-OH, and C14-OH), dodecanoylcarnitine (C12), 7-methylguanine, and sphinganine 1-phosphate [10]. The recurrence of these metabolites and pathways across the expanded cohort supports that the insulin-associated signature reflects a stable biological response to systemic insulin excess rather than cohort-specific artefacts. Thus, providing support for the potential utility of postmortem metabolomics when direct insulin measurement is unreliable.

Beyond classification performance, the metabolite classes contributing to the insulin-associated signature were biologically consistent with known metabolic effects of excess insulin. Acute hyperinsulinaemia induces hypoglycaemia and suppresses lipolysis and fatty acid oxidation, resulting in reduced mitochondrial β-oxidation and a shift away from lipid-derived energy substrates [12,13]. Consistent with reduced fatty acid flux, multiple acylcarnitines were decreased in abundance in insulin intoxication cases, an observation previously seen in insulin-stimulating conditions [12,14,15]. Conversely, relative increases in the abundances of purine/nucleoside and membrane-associated lipid metabolites were observed, which likely reflect cellular energy depletion and membrane/stress responses secondary to hypoglycaemia. These changes are consistent with ATP breakdown to purine degradation products and with previously reported increases in phosphatidylcholines and sphingomyelins in hypoglycaemia-associated states [16]. Importantly, such observations in metabolite abundances were not confined to isolated compounds but occurred consistently across distinct metabolite features, indicate a coordinated metabolic phenotype.

The study employed untargeted HRMS with putative metabolite annotations [17], therefore interpretation herein is made at the metabolite class level rather than at individual molecular identities. Nevertheless, the reproducible and biologically coherent behaviours of these metabolite classes across the training and temporally distinct validation environments strengthens confidence that the observed insulin-associated signature reflects genuine physiological changes related to insulin intoxication.

Overall, these findings suggest that postmortem metabolomics captures downstream systemic consequences of insulin excess, providing an indirect but biologically plausible marker of hypoglycaemia that may complement conventional toxicological analyses when direct insulin measurements are unreliable.

From a forensic perspective, the present findings suggest that postmortem metabolomics may provide complementary biochemical evidence in suspected insulin intoxication cases by capturing downstream systemic metabolic consequences of insulin excess rather than relying on direct insulin quantification. The detection of insulin intoxication after death remains challenging, as postmortem insulin measurements are often unreliable due to rapid degradation, haemolysis, and postmortem redistribution [2]. Consequently, absence of detectable insulin does not necessarily exclude insulin-induced hypoglycaemia, and forensic interpretation may rely heavily on circumstantial evidence or indirect biochemical evidence.

Importantly, the insulin-associated signature demonstrated consistent performance across both a controlled matched cohort and a heterogenous forensic cohort reflective of routine casework. This supports that the observed signal represents a reproducible biological response persisting across temporal validation and varying cohort compositions, rather than a cohort- or model-specific artifact. Within the applicable domain, no false negatives were observed in either test cohort, resulting in high NPV within the studied populations. Collectively, these findings suggest that absence of the metabolomic signature may support exclusion of insulin intoxication in cases where conventional biochemical findings are inconclusive.

Rather than replacing established toxicology methods, metabolomics may provide complimentary biochemical evidence reflecting systemic effects of drug exposures [18]. Given that HRMS is increasingly integrated and needed routine postmortem toxicological screening [19], implementation of such analyses could be achieved with minimal to no additional sample requirements. Together, these findings support the potential role of postmortem metabolomics as an adjunctive forensic approach for evaluating suspected insulin intoxication when direct insulin measurements are unreliable or unavailable.

Several methodological strengths and limitations should be considered when interpreting the present findings. The primary aim of establishing a robust analytical framework for characterising an insulin-associated metabolomic signature is supported by several strengths. First, cases were derived from a national forensic cohort encompassing all of Sweden, with routine toxicological screening performed at a single national laboratory. This reduces centre-specific analytical variability and is reflective of real-word casework in Sweden. Second, the use of temporally distinct training and validation cohorts provides a more rigorous assessment of generalisability than internal cross-validation alone, minimising the risk of overfitting and data leakage [11]. Third, model performance was evaluated both in matched case-control comparisons and in a heterogenous forensic cohort, enabling assessment across differing background compositions and biological variability. Importantly, although predictions were restricted to the DModX-defined applicability domain to minimise extrapolation beyond the validated training space, insulin intoxication cases remained detectable with 100% sensitivity even without DModX filtering. This suggests that the observed insulin-associated signal is not dependent on the applicable domain restriction, while DModX filtering primarily improves confidence and specificity of classifications.

Several limitations should also be considered. First, the study utilised untargeted HRMS data with metabolite annotation via in-house library matching and public database comparison, equivalent to MSI level 2 and level 3 putative metabolite identification, respectively [17]. Therefore, biological interpretation is limited primarily to metabolite class level rather than confirmed structural identification. Second, postmortem metabolomic profiles can be influenced by factors such as postmortem interval, postmortem degradation, and postmortem redistribution [20–22], which cannot be fully controlled in retrospective forensic cohorts. Herein, matching was applied between groups for training and Test Set 1 cohorts in term of biological sex, age, BMI, and PMI, however, residual confounding effects may remain. Third, definitive biochemical confirmation of insulin intoxication remains inherently challenging postmortem, and some degree of diagnostic uncertainty in retrospective case classification cannot be excluded. Finally, although this cohort represents one of the largest reported series of confirmed insulin intoxications, the absolute number of cases remains modest due to the rarity and diagnostic difficulty of this cause of death.

Despite these limitation, the consistent performance of the insulin-associated metabolomic signature across temporally distinct and heterogenous validation settings supports the presence of a reproducible biological signal associated with insulin-induced hypoglycaemia. Further external validation across independent laboratories, analytical platforms, and forensic populations will be necessary to determine broader generalisability and potential forensic applicability.

## CONCLUSIONS

In conclusion, we demonstrate that postmortem metabolomics can identify a reproducible metabolomic signature associated with insulin intoxications across a national forensic cohort and temporally distinct validation settings. The insulin-associated signature remained consistent across both matched and heterogeneous postmortem populations, supporting the presence of a stable biological signal rather than cohort-specific artefacts. The observed metabolite class alterations were biologically consistent with systemic effects of insulin-induced hypoglycaemia, providing physiological plausibility to the detected insulin-associated signature.

Collectively, these findings suggest that untargeted postmortem metabolomics may provide complementary biochemical evidence in investigations of suspected insulin intoxication, particularly when direct insulin measurements are unreliable or inconclusive. While not suitable as a standalone confirmatory diagnostic approach, the consistently high sensitivity and absence of false negatives within the validated applicability domain suggest potential utility in supporting exclusion of insulin intoxication in forensic death investigations. Further validation across independent laboratories and forensic populations will be necessary to determine broader generalisability and forensic applicability

## Data Availability

Data availability
The data that support the findings of this study are available on request from the corresponding author. The data are not publicly available, due to legal and ethical considerations.
Code availability
Code used for data preprocessing, analysis, and visualisations are available at GitHub: github.com/LJ-Ward/insulin.

https://www.github.com/LJ-Ward/insulin

## ADDITIONAL INFORMATION

### Ethics approval and consent to participate

The study was approved by the Swedish Ethical Review Authority (Dnr 2019-04530, 2024-05087-02, 2025-02459-02). Due to the retrospective nature of the study, the need for informed consent was waived by the Swedish Ethical Review Authority. All methods were carried out in accordance with relevant guidelines and regulations.

### Availability of data and material

The processed untargeted metabolomics data that support the findings of this study are available on request from the corresponding author. The data are not publicly available, due to legal and ethical considerations. Code used for data preprocessing, analysis, and visualisations are available at GitHub: github.com/LJ-Ward/insulin.

### Author contributions statement

Conceptualisation, A.E, C.S., H.G., F.C.K and L.J.W.; methodology, A.E. and L.J.W.; software, A.E. and L.J.W.; formal analysis, L.J.W; investigation, A.E., C.S., F.T., H.G., F.C.K. and L.J.W; visualisation, L.J.W.; writing—original draft, L.J.W.; writing—review and editing, A.E., C.S., F.T., H.G., F.C.K. and L.J.W.; All authors have read and agreed to the published version of the manuscript.

