## Supplement Figure S1 for "Untargeted metabolomics reveals a reproducible signature of fatal insulin intoxication"

__List of supplements:__

**Supplemental Table S1:** Validation performance in temporally distinct test sets – hyperglycaemic (hyper) and non-insulin (control) groups.

**Supplemental Table S2:** Validation performance in temporally distinct test sets – without the inclusion of DModX applicable domain filter.

**Supplemental Table S3:** Metabolite annotation of 91 chromatographic features identified as insulin-associated. Annotation yielded 32 features corresponding to 27 unique metabolites; remaining features did not yield confident database matches.

**Supplemental Figure S1:** Permutations testing of OPLS-DA models. Permutation plots generated using 500 random class permutations for the OPLS-DA models comparing (a) insulin intoxication and control cases, (b) insulin intoxication and hyperglycaemic cases, and (c) the three-group model reconstructed following SUS-plot feature selection (see Figure 2). In all models, the original models demonstrated higher Q2 values than permuted models, with negative Q2 intercepts indicating absence of significant model overfitting.

**Supplemental Table S1:** Validation performance in temporally distinct test sets – hyperglycaemic (hyper) and non-insulin (control) groups.

| | Test Set 1 | Test Set 2 |
|---|---|---|
| **Sensitivity – hyper** | 33.3 % | - |
| **Specificity – hyper** | 100 % | - |
| **PPV – hyper** | 100 % | - |
| **NPV – hyper** | 90.2 % | - |
| **Sensitivity – control** | 76.9 % | 72.4 % |
| **Specificity – control** | 88.2 % | 100 % |
| **PPV – control** | 90.9 % | 100 % |
| **NPV – control** | 71.4 % | 30.8 % |

Test Set 1: matched validation cohort reflecting the training set design; Test Set 2: heterogenous validation cohort including insulin cases and randomly selected forensic autopsy cases.
Sensitivity, specificity, positive predictive value (PPV), negative predictive value (NPV), and accuracy calculated among cases within the distance to model in X-space (DModX)-defined applicability domain.

**Supplemental Table S2:** Validation performance in temporally distinct test sets – without the inclusion of DModX applicable domain filter.

| | **Test Set 1** | **Test Set 2** |
|---|---|---|
| **Sensitivity – insulin** | 100 % | 100 % |
| **Specificity – insulin** | 66.7 % | 65.0 % |
| **PPV – insulin** | 51.9 % | 22.2 % |
| **NPV – insulin** | 100 % | 100 % |
| **Accuracy** | 71.2 % | 68.4 % |

Test Set 1: matched validation cohort reflecting the training set design; Test Set 2: heterogenous validation cohort including insulin cases and randomly selected forensic autopsy cases.

**Supplemental Table S3:** Metabolite annotation of 91 chromatographic features identified as insulin-associated. Annotation yielded 32 features corresponding to 27 unique metabolites; remaining features did not yield confident database matches.

| m/z | Rt (s) | Identification† | log2FC (insulin/control) | Adjusted p-value (FDR) |
|---|---|---|---|---|
| 303.2320 | 626.54 | Eicosapentaenoic acid* | 1.08 | 6.92E-04 |
| 738.5104 | 614.13 | | 0.98 | 2.35E-04 |
| 739.5178 | 619.08 | | 0.91 | 5.04E-04 |
| 304.2353 | 626.74 | Eicosapentaenoic acid* | 0.78 | 9.34E-03 |
| 204.0281 | 78.45 | | 0.76 | 1.67E-02 |
| 588.3660 | 585.85 | | 0.75 | 7.22E-03 |
| 307.2231 | 51.29 | 3-hydroxyheptanoylcarnitine | 0.73 | 1.98E-03 |
| 326.1764 | 599.68 | | 0.72 | 8.96E-04 |
| 464.3374 | 577.05 | | 0.69 | 8.18E-04 |
| 303.3087 | 617.93 | | 0.69 | 2.69E-04 |
| 244.1667 | 50.67 | Lysylproline | 0.67 | 4.48E-03 |
| 161.1367 | 60.21 | | 0.63 | 5.61E-03 |
| 449.3456 | 624.70 | | 0.63 | 5.04E-05 |
| 791.6271 | 641.81 | | 0.62 | 9.34E-03 |
| 263.0747 | 135.81 | | 0.60 | 5.04E-04 |
| 475.3617 | 632.51 | | 0.56 | 2.95E-03 |
| 204.0282 | 118.33 | | 0.53 | 2.67E-03 |
| 472.3424 | 622.59 | Cervonyl carnitine | 0.52 | 7.22E-03 |
| 473.3457 | 621.98 | | 0.48 | 2.57E-03 |
| 279.0491 | 135.34 | | 0.48 | 5.61E-03 |
| 116.1069 | 57.26 | 2-Hexenal | 0.48 | 3.46E-02 |
| 465.3418 | 574.19 | | 0.46 | 2.75E-03 |
| 188.0542 | 118.35 | 7-Methylguanine | 0.44 | 3.39E-03 |
| 477.3769 | 647.93 | | 0.44 | 2.93E-01 |
| 208.1624 | 363.63 | | 0.43 | 1.62E-02 |
| 301.2164 | 614.48 | all-trans-Retinoic acid* | 0.42 | 3.73E-02 |
| 447.3301 | 609.86 | | 0.41 | 2.25E-02 |
| 739.5162 | 641.53 | SM(34:2) | 0.4 | 6.24E-02 |
| 478.3890 | 648.70 | | 0.38 | 5.98E-02 |
| 291.2531 | 619.66 | | 0.37 | 1.69E-01 |
| 151.0614 | 77.39 | Methylhypoxanthine | 0.34 | 1.63E-01 |
| 148.0427 | 74.35 | Methionine sulfoxide | 0.32 | 8.88E-02 |
| 431.1536 | 77.89 | | 0.29 | 4.27E-01 |
| 382.2738 | 595.18 | | 0.26 | 1.20E-01 |
| 141.1135 | 618.47 | | 0.24 | 4.06E-01 |
| 330.1029 | 375.97 | | 0.20 | 1.48E-01 |
| 166.0722 | 118.47 | 7-Methylguanine | 0.18 | 3.48E-01 |
| 712.5300 | 647.12 | | 0.17 | 4.19E-01 |
| 394.1569 | 75.75 | | 0.17 | 5.63E-01 |
| 850.8450 | 391.20 | | 0.17 | 5.63E-01 |
| 158.1173 | 74.17 | | 0.09 | 5.63E-01 |
| 301.2931 | 608.25 | | 0.03 | 9.14E-01 |
| 254.1507 | 49.62 | | 0.01 | 9.42E-01 |
| 916.6330 | 417.19 | | -0.02 | 9.42E-01 |
| 926.6373 | 416.36 | | -0.02 | 9.42E-01 |
| 428.3369 | 652.87 | | -0.04 | 8.98E-01 |
| 281.0743 | 133.20 | Ribothymidine | -0.05 | 8.66E-01 |
| 916.9664 | 417.23 | | -0.05 | 8.87E-01 |
| 276.1807 | 144.22 | | -0.08 | 8.66E-01 |
| 404.2555 | 595.21 | Sphinganine 1-phosphate | -0.10 | 5.63E-01 |
| 300.3259 | 609.98 | | -0.16 | 7.21E-01 |

| | | | | |
|---|---|---|---|---|
| 115.0947 | 197.08 | | -0.20 | 2.93E-01 |
| 546.2626 | 606.20 | | -0.22 | 2.57E-01 |
| 371.2992 | 588.58 | | -0.23 | 4.19E-01 |
| 219.0265 | 45.27 | | -0.29 | 2.93E-01 |
| 140.0682 | 46.58 | L-Valine* | -0.31 | 1.10E-01 |
| 274.2011 | 329.15 | Heptanoyl carnitine | -0.33 | 1.85E-01 |
| 248.1493 | 116.99 | 3-Hydroxybutyrylcarnitine* | -0.33 | 4.41E-02 |
| 458.3476 | 575.01 | | -0.35 | 1.89E-01 |
| 269.1008 | 61.23 | | -0.37 | 9.34E-03 |
| 231.0836 | 133.16 | | -0.38 | 1.10E-01 |
| 114.0549 | 61.83 | 1-Pyrroline-2-carboxylate | -0.39 | 2.93E-01 |
| 137.0711 | 49.63 | 1-Methylnicotinamide* | -0.41 | 3.03E-01 |
| 366.2645 | 570.00 | 3-Hydroxy-5,8-tetradecadienoylcarnitine | -0.44 | 5.61E-03 |
| 360.2748 | 499.09 | 3-Hydroxydodecanoyl carnitine* | -0.44 | 2.11E-02 |
| 388.3056 | 568.33 | 3-Hydroxytetradecanoyl carnitine | -0.48 | 1.18E-02 |
| 286.1064 | 75.66 | 3-Hydroxybutyrylcarnitine* | -0.50 | 5.27E-02 |
| 344.2801 | 570.85 | Dodecanoylcarnitine* | -0.50 | 2.29E-01 |
| 384.3111 | 606.55 | 3-hydroxypentadecanoyl carnitine | -0.53 | 2.48E-03 |
| 443.0827 | 62.18 | | -0.54 | 2.29E-02 |
| 441.3419 | 596.09 | | -0.55 | 7.94E-03 |
| 345.2827 | 570.35 | Dodecanoylcarnitine* | -0.55 | 2.60E-02 |
| 291.0696 | 62.81 | Inosine* | -0.61 | 8.96E-04 |
| 299.0976 | 206.41 | | -0.63 | 6.41E-03 |
| 402.2851 | 458.24 | | -0.63 | 7.94E-03 |
| 389.3094 | 568.70 | 3-Hydroxytetradecanoyl carnitine | -0.63 | 8.18E-04 |
| 193.1582 | 451.49 | | -0.63 | 1.33E-01 |
| 321.0627 | 186.65 | | -0.64 | 2.95E-03 |
| 306.2172 | 219.05 | | -0.64 | 2.94E-02 |
| 343.2668 | 541.89 | | -0.65 | 3.91E-02 |
| 444.3108 | 398.68 | | -0.71 | 6.24E-02 |
| 385.1283 | 61.57 | S-Adenosylhomocysteine | -0.74 | 8.18E-04 |
| 337.0367 | 186.65 | | -0.76 | 2.95E-03 |
| 299.0818 | 186.78 | | -0.81 | 5.04E-05 |
| 260.1856 | 268.00 | Hexanoylcarnitine* | -0.85 | 8.96E-04 |
| 322.0658 | 186.79 | | -0.86 | 2.69E-04 |
| 287.2441 | 59.22 | N1,N12-Diacetylspermine | -0.94 | 5.04E-05 |
| 163.0422 | 186.75 | | -0.97 | 5.04E-05 |
| 374.2536 | 384.49 | Dodecanedioylcarnitine | -1.03 | 2.71E-03 |
| 184.0303 | 65.45 | | -1.16 | 5.04E-05 |
| 227.0291 | 159.25 | | -1.33 | 5.04E-05 |

* Metabolite annotation by accurate mass matching to internal reference library with retention time confirmation where available.

† Blank identification entries indicate features without confident metabolite annotation when matching against an internal reference library, or by accurate mass matching to public databases (HMDB) for features without internal matches.

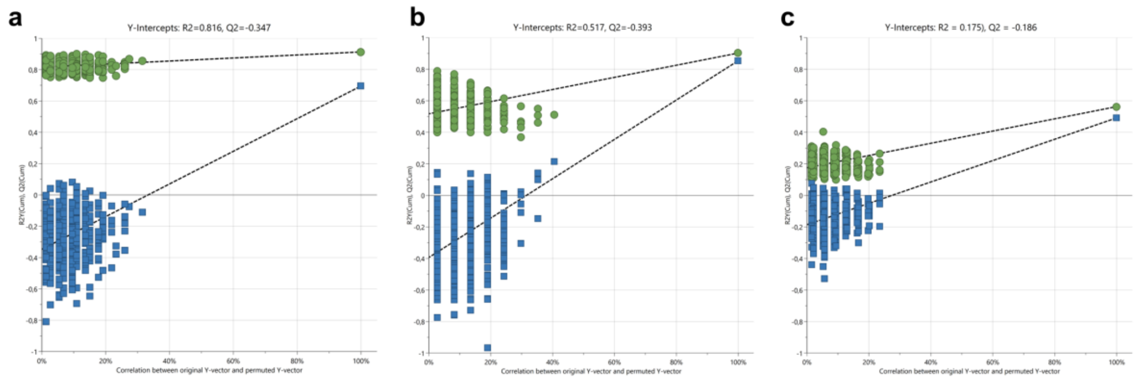

Supplemental Figure S1: Permutations testing of OPLS-DA models. Permutation plots generated using 500 random class permutations for the OPLS-DA models comparing (a) insulin intoxication and control cases, (b) insulin intoxication and hyperglycaemic cases, and (c) the three-group model reconstructed following SUS-plot feature selection (see Figure 2). In all models, the original models demonstrated higher Q2 values than permuted models, with negative Q2 intercepts indicating absence of significant model overfitting.
